# Analysis and Prediction of the COVID-19 outbreak in Pakistan

**DOI:** 10.1101/2020.06.21.20136341

**Authors:** Mohsin Ali, Mudassar Imran, Adnan Khan

**Author notes:** Data data availability statement : The data that support the findings of this study are openly available in at COVID-19 Data Repository by the Center for Systems Science and Engineering (CSSE) at Johns Hopkins University [https://github.com/CSSEGISandData/COVID-19], reference number [32].

## Abstract

In this study we estimate the severity of the COVID-19 outbreak in Pakistan prior to and after lock down restrictions were eased. We also project the epidemic curve considering realistic quarantine, social distancing and possible medication scenarios. We use a deterministic epidemic model that includes asymptomatic, quarantined, isolated and medicated population compartments for our analysis. We calculate the basic reproduction number ℛ_0_ for the pre and post lock down periods, noting that during this time no medication was available.^1^ The pre-lock down value of ℛ_0_ is estimated to be 1.07 and the post lock down value is estimated to be 1.86. We use this analysis to project the epidemic curve for a variety of lock down, social distancing and medication scenarios. We note that if no substantial efforts are made to contain the epidemic, it will peak in mid of September, with the maximum projected active cases being close to 700,000. In a realistic, best case scenario, we project that the epidemic peaks in early to mid July with the maximum active cases being around 120000.We note that social distancing measures and medication if available will help flatten the curve, however without the reintroduction of further lock down it would be very difficult to bring ℛ_0_ below 1. Our study strongly supports the recent WHO recommendation of reintroducing lock downs to control the epidemic.

## 1 Introduction

Coronavirus disease (COVID-19) is a respiratory tract illness which originated in Wuhan, China in early December 2019. It spread to other countries in Asia, Europe and North America in early 2020 and was declared a pandemic by the WHO on March 11 2020. As of June 15 2020, the disease is prevalent in at least 212 countries and territories, with more than 8 million cases reported and around 435,000 fatalities [3].

COVID-19 is caused by a coronavirus SARS-CoV-2 [2], belonging to a family of viruses that are found in humans and different species of animals including cattle, camels and bats. These have in the past caused serious disease outbreaks in human populations as happened in MERS (caused by MERS-CoV) and SARS (caused by SARS-CoV) [1, 4, 5].

The virus spreads primarily from person to person via through respiratory droplets [1]. Symptoms of the disease may appear 2-14 days after exposure and may include fever, cough shortness of breath, chills, muscle pain and loss of taste or smell[2, 6]. These may range from very mild (in 80 % of the cases) to severe (in 15 % of the cases) to critical (in 5 % of the cases)[2]. Those at higher risk for severe illness include the elderly (people ages 65 and above) and people with co morbidities such as chronic lung diseases, diabetes, chronic kidney diseases, serious heart conditions and immunocompromised individuals[7].

At this time there is no approved vaccine for COVID-19 although some possible possible vaccines are undergoing Clinical trials [12]. Several antiviral drugs are now in advanced clinical trial phase [8, 9, 10, 14, 13]. The trials for these drugs have so far have produced mixed results with some drug regimens showing encouraging outcomes, such as shortening of the duration of the disease, while others showing no significant change in the outcomes due to medication. A drug that has been suggested to have beneficial effects in COVID-19 treatment is Remdesivir, in their study Grein et.al [14] have reported improvement in 68% of the patients in a controlled trial using Remdesivir, similarly Beigel et.al [8] have also reported shortening in the time to recovery with Remdesivir use. In another study involving triple anti viral therapy Huang et.al [6] the authors have also reported alleviation of symptoms and shortening the duration of the disease. At the time of writing Remdesivir has received emergency approval in a few countries [11].

Non pharmaceutical interventions such as social distancing measures, isolation and quarantine had a positive effect in controlling the outbreak, however these measures specially lock downs also resulted in substantial economic and social cost [15] and as a consequence countries are easing the lock downs. For this reason, in the near future, social distancing and hopefully the availability of medication soon, are the viable strategies that can be used to control the epidemic.

After the initial outbreak in China, the COVID-19 spread world wide with Europe becoming the epicenter of the outbreak [2], followed by the US. South Asia, specifically India and Pakistan while also affected, with cases being reported in early March 2020, have had a very different epidemic curve as compared to China, Europe and North America, with a much lower disease burden and mortality rate. There has been much speculation as to why the outbreak has been so different in these countries, from the more mundane theories citing effective and early quarantine and isolation, demographics and higher temperatures to more esoteric ones that hold a better immune response or previous vaccinations (which are no longer required in the West) as possible reasons. However to our knowledge no proper study has been undertaken and all of these are conjectures at the moment. As these countries have eased the lock down, the COVID-19 cases have started to increase at a high rate, the morbidity though still remains low as compared to other countries [16].

Since the outbreak many studies have been undertaken to estimate the growth rates and understand the transmission dynamics of COVID-19. These include phenomenological models[17, 18], stochastic models[19], both of which are very useful in the early stages of the outbreak, and mechanistic models[20, 21, 22, 23, 25] that incorporate our understanding of the transmission pathways. Imran et.al [24] and Perkins et.al [26] have used optimal control techniques to propose efficient control strategies. Gumel et.al [23] have considered the impact of various non pharmaceutical interventions on the disease burden and mortality. The aim of such modeling is twofold, one to provide estimates of the severity of the outbreak by calculating quantities like the growth trends of the epidemic, estimates of the final outbreak size and duration of the outbreak and second to provide insights into efficacy of various control measures.

The COVID-19 epidemic in Pakistan started in early March 2020, with the initial cases being reported in late February 2020. Lock downs that were imposed in different parts of the country in the third week of March in order to control the outbreak were successful in keeping the disease numbers low. However with the start of the muslim holy month of Ramadan on 24 April, the lock down measure became less effective and by May 1 the lock down restriction were officially eased with complete relaxation on 22 May. As expected this lead to an upsurge in the infections with close to 80,000 active cases and around 132000 total cases being reported by June 12. Considering the situation on 10 June the WHO issued an advisory strongly recommending an intermittent lock down for the following two weeks. In this study we model the COVID 19 outbreak using a variant of the model proposed by Imran et.al [24] for the transmission dynamics of the disease. This model incorporates compartments for quarantined, isolated, and asymptomatic individuals, pathways considered important in transmission of the disease. We also include a compartment for individuals taking medication as we would like to study the possible effects of medication in controlling the disease. The dynamics of the COVID 19 outbreak in Pakistan have been qualitatively different before and after the lock down was eased. We estimate the value of ℛ_0_ in Pakistan for the two different phases of the outbreak, noting that during this time no medication was available. We then project the disease curve considering minimal intervention as well as various control strategies. At the moment the viable strategies for controlling the outbreak are quarantine, isolation, social distancing and possibly medication. In fact, there have been emergency approvals for different treatments for the disease in various countries including Pakistan. We project the epidemic curve considering various quarantine, social distancing and medication scenarios. We note that the most effective control measure is a lock down but a very strict lock down does not seem to be an option that is being considered by the government, however a moderate and intermittent lock down may be possible. Further, by making sure social distancing protocols issued by the government are followed we can reduce the contact rate to varying degrees also flattening the disease curve. Finally, we consider various medication scenarios, as mentioned medication to date has been shown in varying degrees, to reduce the time of infection as well as alleviating symptoms, we primarily consider medication in the context of shortening the time of infection and thereby helping to bring the disease numbers down. We look at various levels of quarantine, social distancing and medication to study how each of these would affect the disease numbers as well as when would the infected numbers peak under them. We conclude the study by summarizing our findings and suggesting realistic control measures with their impact on the epidemic.

## 2 Model Formulation

The COVID-19 transmission model we consider is based upon the SEIR model and takes into account the effects of quarantine, isolation, medication, and asymptomatic individuals. The total population *N* (*t*) is divided into eight mutually exclusive subpopulations, susceptibles *S*, these are individuals who can fall ill by coming in contact with an infected individual, quarantined susceptibles *Q*_*S*_, these individuals are removed from the susceptible group at rate *ϵ*, either through self quarantined or lock-down measures, they, however, go back to the susceptible group at rate *ξ*. The susceptibles move to the exposed class by coming in contact with any infectious individual, at rate *ρλ*, some exposed individuals will not show symptoms and are accounted for in the model by the movement to the asymptomatic class at rate (1 *− ρ*)*λ*. Exposed individuals *E* become infected at rate *σ*. Infected individuals *I* can be isolated or given medication at a rate of *τ* and *α*_*I*_ whereas a fraction of isolated *Q*_*I*_ are given medication at rate *α*_*Q*_. The individuals taking medication are represented by a separate compartment *M*, these include individuals from the isolated and infected classes who are under treatment. Based on clinical trials thus far, the main effect of medication we incorporate is that, it shortens the duration of the disease [6, 8, 14]. The recovery time for infected *I*, asymptomatic *A*, isolation *Q*_*I*_ and medication *M* is given by 1*/γ*,1*/θ*_*A*_,1*/θ* and 1*/θ*_*M*_ respectively. The schematic of the transmission pathways is given in Fig.1. The total population *N* (*t*) is given by the sum of the sub-populations.

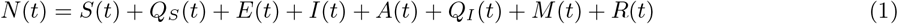

The governing equations are given below (2).

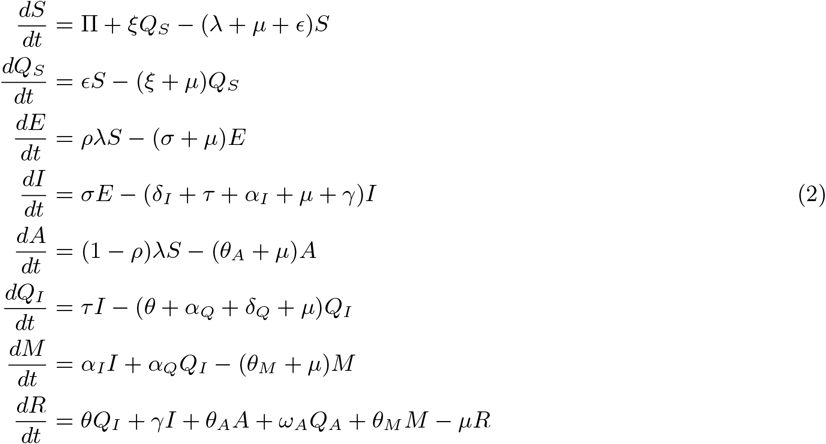

where *λ* is the force of infection

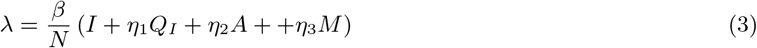

Here *β* is the effective contact rate, where as *η*_1_,*η*_2_ and *η*_4_ are the associated relative infectiousness parameters for the *Q*_*I*_, *A* and *M* sub-populations. Table.2 and Table.3 represent the description of variables and parameters of the model, and are given in the appendix.

## 3 Basic Properties

The state variables described in model (2) shows non-negative solutions for all time *t ≥* 0 with non-negative initial conditions.

### Lemma 3.1.

*For any given non-negative initial conditions, there exist a unique solution*

*S, Q*_*S*_, *E, I, A, Q*_*I*_, *M, R respectively, for all t ≥* 0. *Moreover, it satisfy the following inequality of boundedness*.

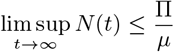

Proof is presented in the appendix A.

### Lemma 3.2.

*The closed set:*

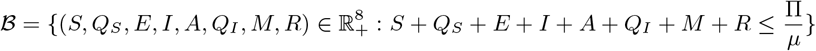

*is positively invariant*.

Proof is attached in the appendix A.

## 4 Steady State Analysis

### 4.1 Disease Free Equilibrium

The model (2) achieves the Disease Free Equilibrium (DFE) whenever there is no induction of infection by the disease i.e. the force of infection is zero,*λ* = 0. Mathematically, this can be donr by equating the right hand side of (2) to zero with *λ* = 0. Let *F*_0_ represent the DFE of the model.

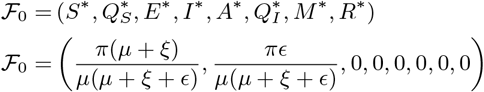

The local stability of Disease free equilibrium (DFE) is quantified by the threshold quantity ℛ_0_ which is found by means of the next generation operator method [27].

### 4.2 The Basic Reproduction Number ℛ_0_

The thresh hold quantity ℛ represents the average number of new secondary infections produced by the single infection in the completely susceptible population. It is calculated by the spectral radius of the *FV* ^*−*1^ matrix from the next generation method [27]. The associated *F* and *V* matrices of model (2) are as follows

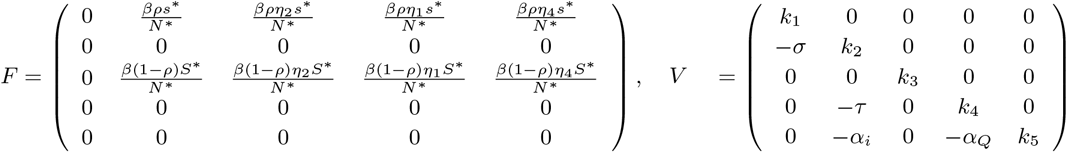

where *k*_1_ = *κ*_*E*_ + *σ* + *µ, k*_2_ = *δ*_*I*_ + *τ* + *γ* + *α*_*I*_ + *µ, k*_3_ = *θ*_*A*_ + *µ, k*_4_ = *θ* + *δ*_*Q*_ + *α*_*Q*_ + *µ, k*_5_ = *θ*_*M*_ + *µ*

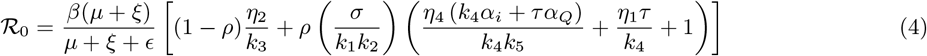

#### Stability of DFE

##### Lemma 4.1.

*The steady state (DFE)* ℱ_0_ *of the model* (2) *is locally-asymptotically stable if* ℛ_0_ *<* 1, *and unstable if* ℛ_0_ *>* 1.

This Lemma interprets that a small influx of the infecties will not lead to cause bigger outbreaks and disease infection will become extinct in long run.

To have concrete sense, that the disease extinction is independent on the initially sub-populations in model (2), we have to show that the steady state (DFE) follows globally asymptotically stable (GAS) condition.

##### Lemma 4.2.

*The Steady state (DFE) ℱ*_0_ *of model* (2) *is globally asymptotically stable if* ℛ_0_ *<* 1 *and unstable* ℛ_0_ *>* 1, *whenever E* = 0

Proof is given in appendix A

### 4.3 Endemic Equilibrium

The equilibrium state of the system (2) in presence of the infection i.e. *λ ≠* 0 is known as the endemic equilibrium. Let ℱ_1_ represents the arbitrary endemic equilibrium of the model (2).

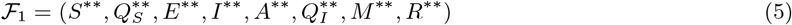

Further,The force of infection *λ* can be written in terms of the equilibrium as

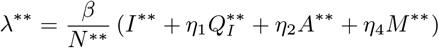

with 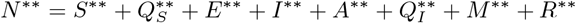

Solving for the system (2) at this specific fixed point, the endemic equilibrium becomes

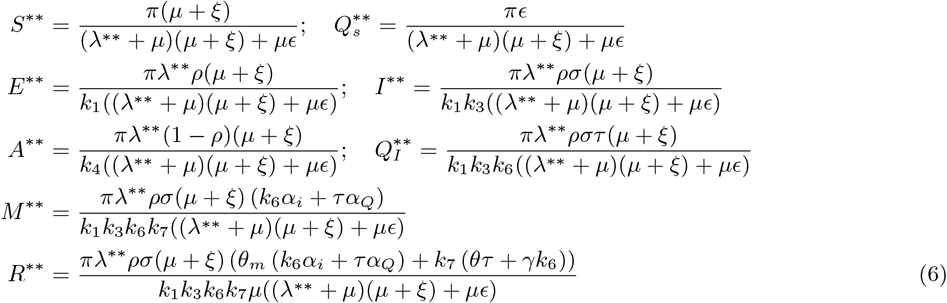

### 4.4 Variation of ℛ_0_ with different Control Parameters

We would like to study how ℛ_0_ changes with the parameters of the model, in particular we are interested in the variation of ℛ_0_ as the quarantine rate, the contact rate and the rate of medication are varied. These rates can be changed by the use of different control strategies, this in turn will be used to project the epidemic curve under various control regimes.

In Figure 2(a) the contours of ℛ_0_ are plotted against *α*_*I*_ the rate of medication and *β* the contact rate. In our analysis we assume that the contact rate can be changed by following stricter social distancing protocols, the medication rate of course can be varied by wider use of available medication. We have also assumed that available medication reduced the duration of the disease by 25%, as reported in several studies. We note that while a higher medication rate (with this particular efficacy) reduces ℛ_0_ medication by itself cannot make ℛ_0_ *<* 1 for high contact rates. Figure 2(b) is a contour plot of ℛ_0_ with quarantine rate (*ϵ*) and the contact rate (*β*). The primary mechanism of quarantine is lock down, although self quarantine can also help. We note that ℛ_0_ can be reduced significantly by either enforcing strict social distancing, thereby reducing *β* or by enforcing a lock down and reducing *ϵ*. In Figure 2(c) contours of ℛ_0_ are plotted against the medication rate and the quarantine rate. We note again that while medication does lower the value of ℛ_0_, for very low quarantine rates just using medication as a control strategy cannot make ℛ_0_ *<* 1.

**Figure 1:**
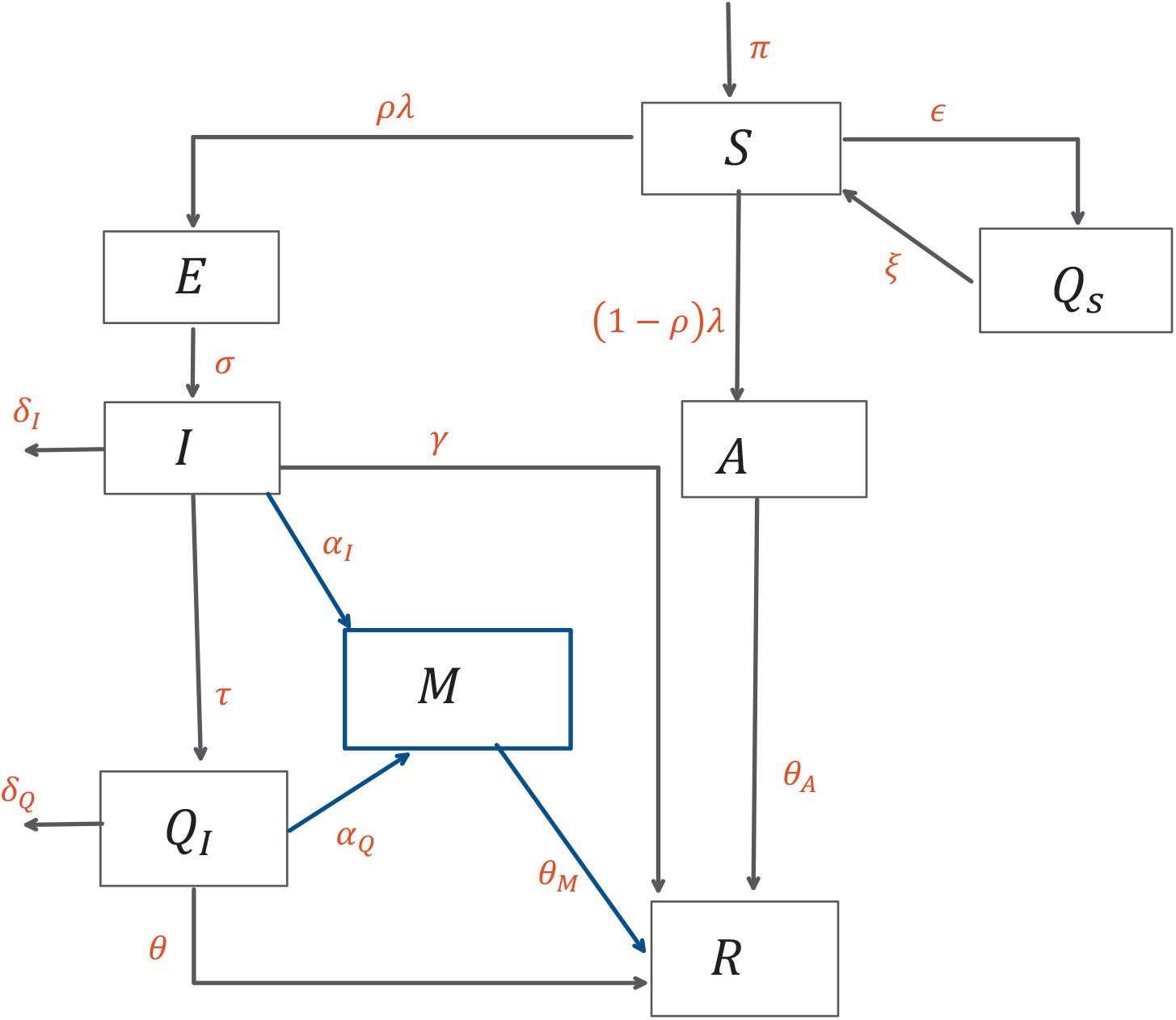
Schematic diagram of the model (2)

**Figure 2:**
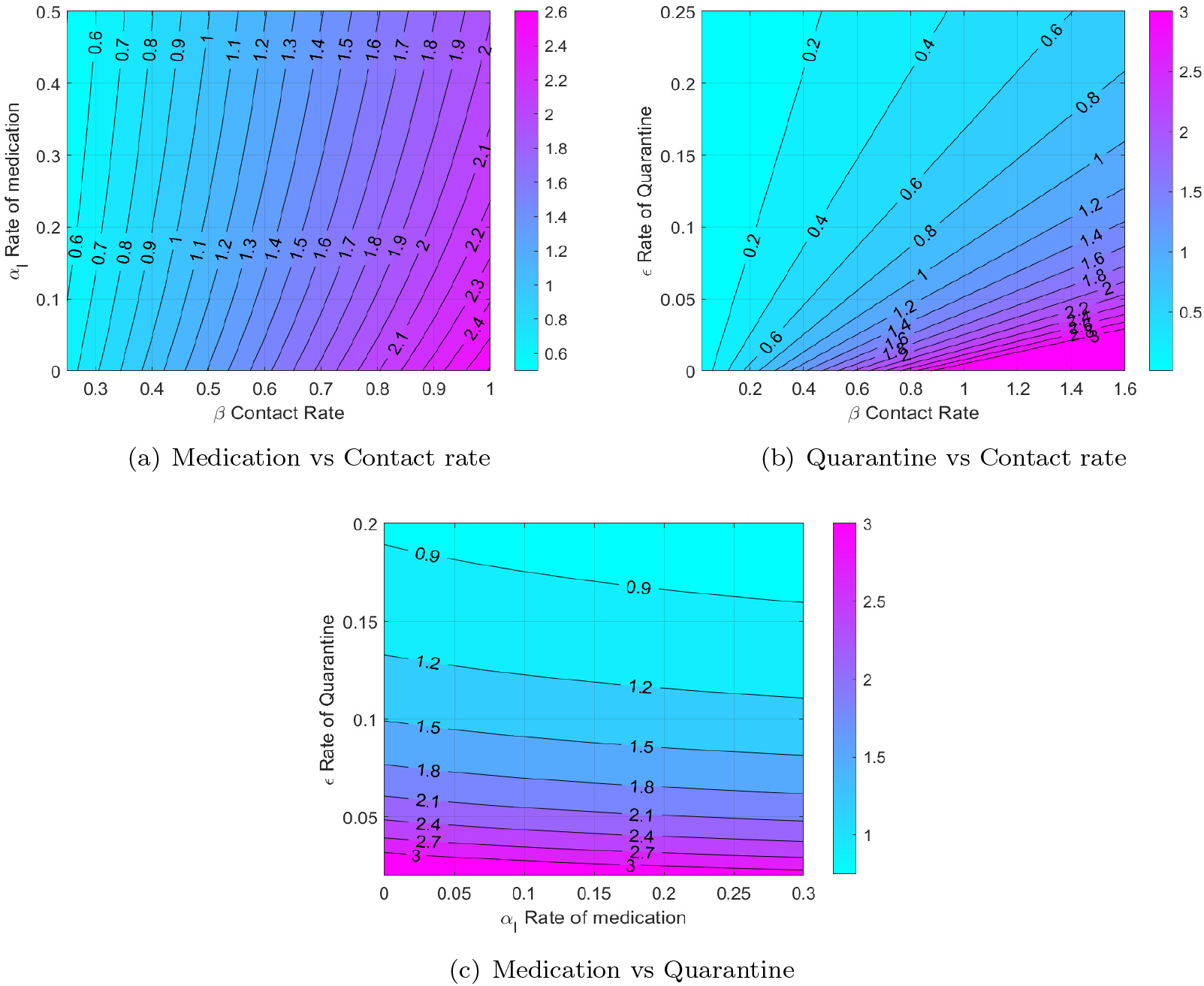
Contour comparison of ℛ_0_

## 5 Estimation and Forecasting

### 5.1 Data sources

The epidemic data of COVID-19 Pakistan for this study is sourced from the COVID-19 Data Repository by the Center for Systems Science and Engineering (CSSE) at Johns Hopkins University [32]. The epidemic data consist of three time series of confirmed cases, deaths and the recovered cases spanning over more than ten weeks till June 15,2020, including the lock down period of 5 weeks. The active cases time series shown in Fig 3 is calculated as

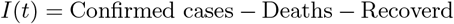

**Figure 3:**
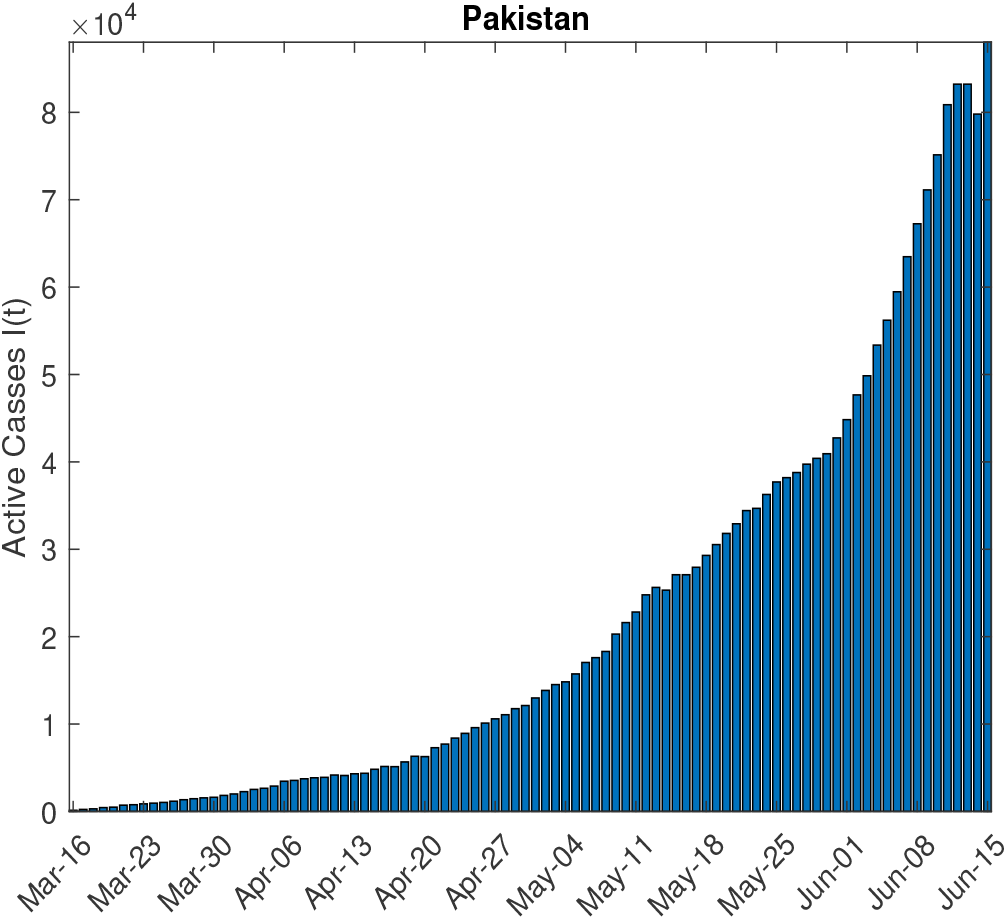
COVID-19 Epidemic data of Pakistan

**Figure 4:**
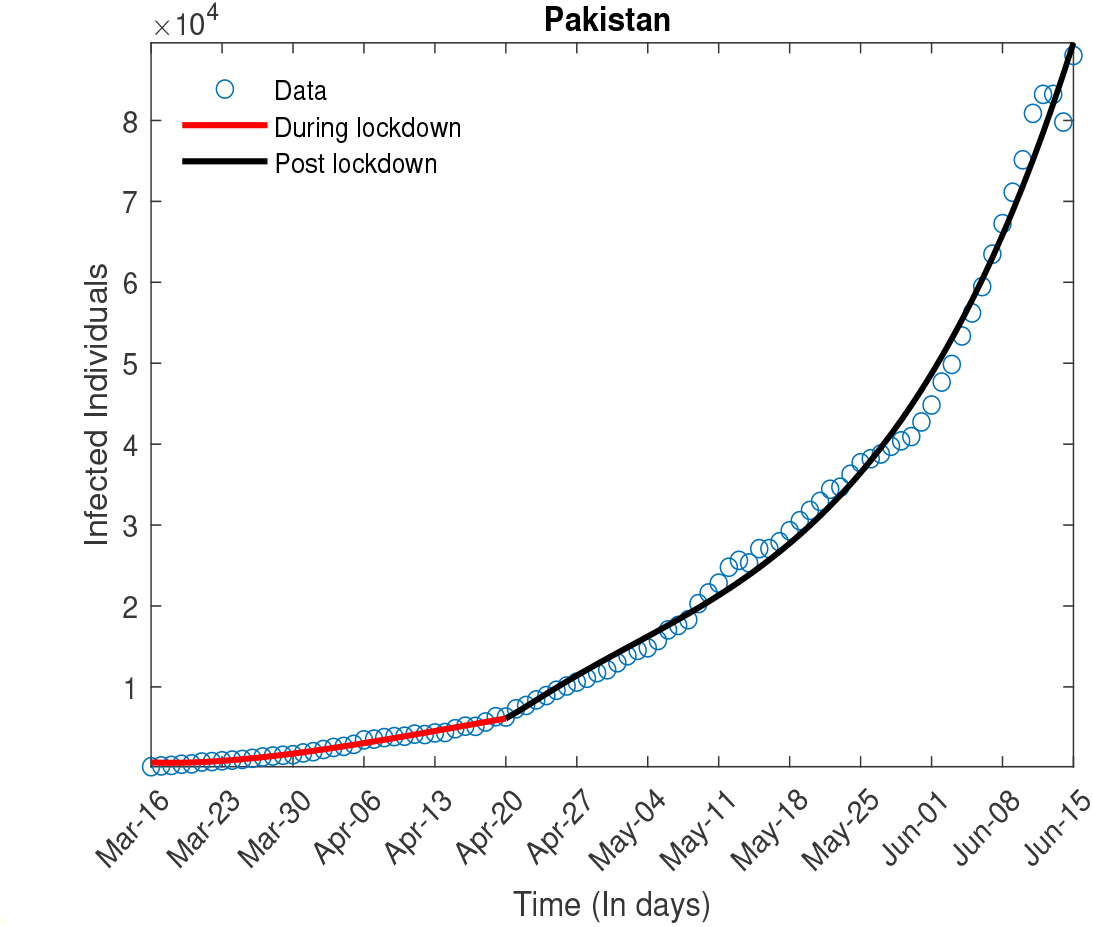
Estimation of ℛ_0_

### 5.2 Estimation

The transmissibility of an infection is quantified by a threshold quantity ℛ_0_,which defined as average number of newly infected individuals by a single infected individual in complete susceptible population. For an ongoing epidemic of COVID-19, Estimates of ℛ_0_ can give better insights about the transmissibility of infection in a certain country. These Estimates further depend upon the critical parameter such as contact rate *β*, incubation rate *σ* and other related parameters. Since there are control polices such as social distancing measures and quarantine being enforced in order to lessen effective contacts between susceptible and infected population groups. It is assumed that the effective contact rate is function of time. During lock down individuals will have less interaction as compared to after lock down so it is assumed to have two different contact rates of *β*, hence giving us a different transmissibility index.

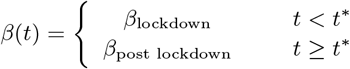

where *t*^*\**^ is the time when lock down was relaxed. For Pakistan, the epidemic curve has followed two very different trajectories before and after the lock down was eased. As we have described in detail in the Introduction the strict lock down was relaxed around the beginning of the fourth week of April, hence *t*^*\**^ is taken to be 5 weeks starting from the March 16,2020.

Since the direct estimates of parameters are really difficult especially, when epidemic/pandemic is still going on. However, we can adopt an indirect method proposed by the [30, 31] to estimates these parameters from the infected data available. The Ordinary Least square optimization technique is used which assumed that the observed data has the constant variance error distribution.

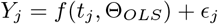

where *Y*_*j*_ are the observed values, Θ_*OLS*_ are set of optimized parameters such as *β, γ, σ* etc using standard OLS optimization (7) And *f* (·) is nonlinear function associated with fitting model. In this case, the model (2) is the underlying model for fitting.

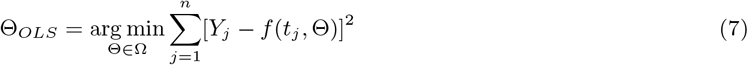

We estimate different parameters of the model, specifically the contact rate *β* is estimated, this in turn is used to determine the value of ℛ_0_ prior to and after the relaxation of the lock down. The values are given in the table below.

**Table 1:** Estimates of ℛ_0_ for Different Epidemic Phases

| Phase time | $\mathcal{R}_0$ | $\beta$ | $\epsilon$ |
| --- | --- | --- | --- |
| During Lock down | 1.07 | 1.39 | 0.172 |
| After Lock down | 1.8614 | 1.77 | 0.08 |

The value of ℛ_0_ post relaxation is significantly higher as can be expected, we will use this to project the epidemic curve under a variety of scenarios. We also note the significant reduction in the quarantine rate once the lock down was relaxed.

### 5.3 Forecasting

In this section we project the epidemic curve under different quarantine, social distancing and medication strategies. At the moment there is no government mandated lock down in Pakistan, however some people are observing self quarantine measures, there are however guidelines for proper social distancing in public spaces, and as the infected numbers rise the health authorities are enforcing those guidelines, we consider this to essentially reduce the effective contact rate, finally medication has been approved by the government to treat COVID 19, and we hope that in the coming days it will become widely available. We consider three possible quarantine scenarios, the first where no additional steps are taken by the government, we call this the ‘minimal quarantine’, the second ‘moderate quarantine’, where some additional lock down measures are enforced to increase the quarantine rate to be twice of the post lock down value and finally a ‘strict quarantine’ where measures are taken to bring the quarantine rate back to the pre lock down levels. For each one of these cases we consider high, medium and low levels of social distancing leading to reduced effective contact rates, both with medication and without the availability of medication.

In Figure 5 we consider the scenario where no steps to lock down are taken and the quarantine rate remains at the post lock down level for different contact rates. Decreasing the contact rate not only lowers the peak of the epidemic curve but also shifts it to the left, which means that that the disease will peak earlier. For the case when no medication is available Figure 5(a) we note that the the peak will be reached in mid to late September with the maximum number of active infected being around 700000 for low levels of social distancing, 400000 for moderate social distancing and 200000 if high levels of social distancing are enforced. We also note that in these scenarios the case related fatalities that can be expected by December 2020 are respectively 2.5, 2 and 1.5 million. In Figure 5(b) for when medication is available we note that the disease peaks between August and early September and the maximum active infected number are now reduced to 275000, 150000 and 110000 in the three different social distancing scenarios. Around 1 million, 700000 and 450000 deaths can occur in these cases by the years end.

**Figure 5:**
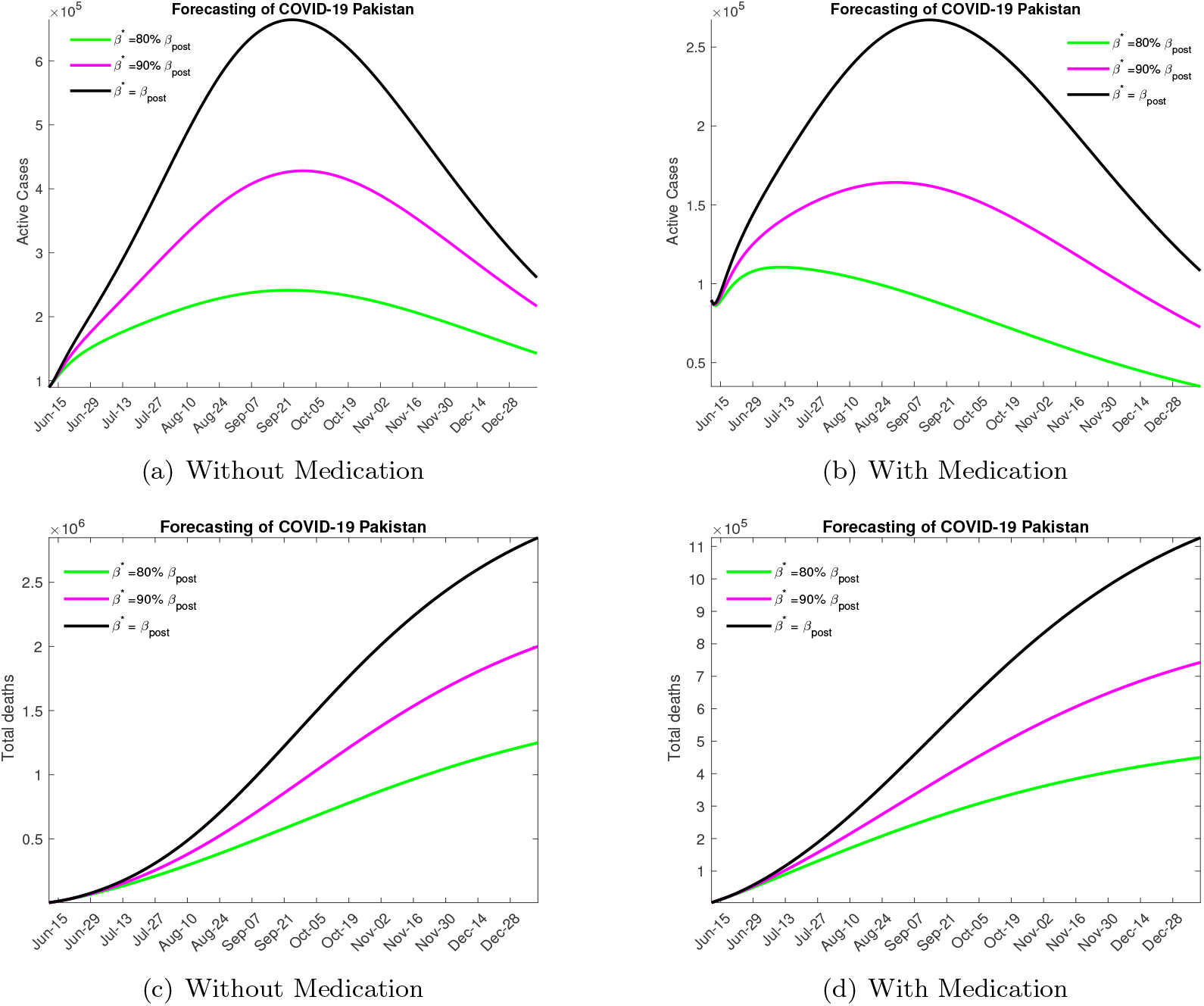
Forecasting with Mild Lock down

We would like to note that this is not a very likely scenario as the rising numbers of infected and disease related deaths have prompted a response both in terms of some additional lock down measures and stricter implementation of the social distancing protocols.

In Figure 6 we consider the outcome when a moderate lock down is imposed increasing the quarantine rate to twice the post lock down level. Without medication Figure 6(a) we note that the the peak will be reached in mid to late July if social distancing protocols are enforced and in mid to late August otherwise, the maximum number of active infected in these cases is projected to be around 150000, 175000 and 300000. The total expected fatalities due to disease are around 500000, 800000 and 1.2 million respectively, through December. In Figure 6(b) for the case medication we note that disease peaks between early to mid July and the maximum active infected number with a high level of social distancing is around 100000, with disease related fatalities through December projected to be around 225000. With moderate social distancing the maximum infected number is projected to be 120000 with 325000 expected deaths by the end of the year. Finally, with low social distancing efforts we expect 150000 active infected cases at the peak of the outbreak and 500000 total fatalities.

**Figure 6:**
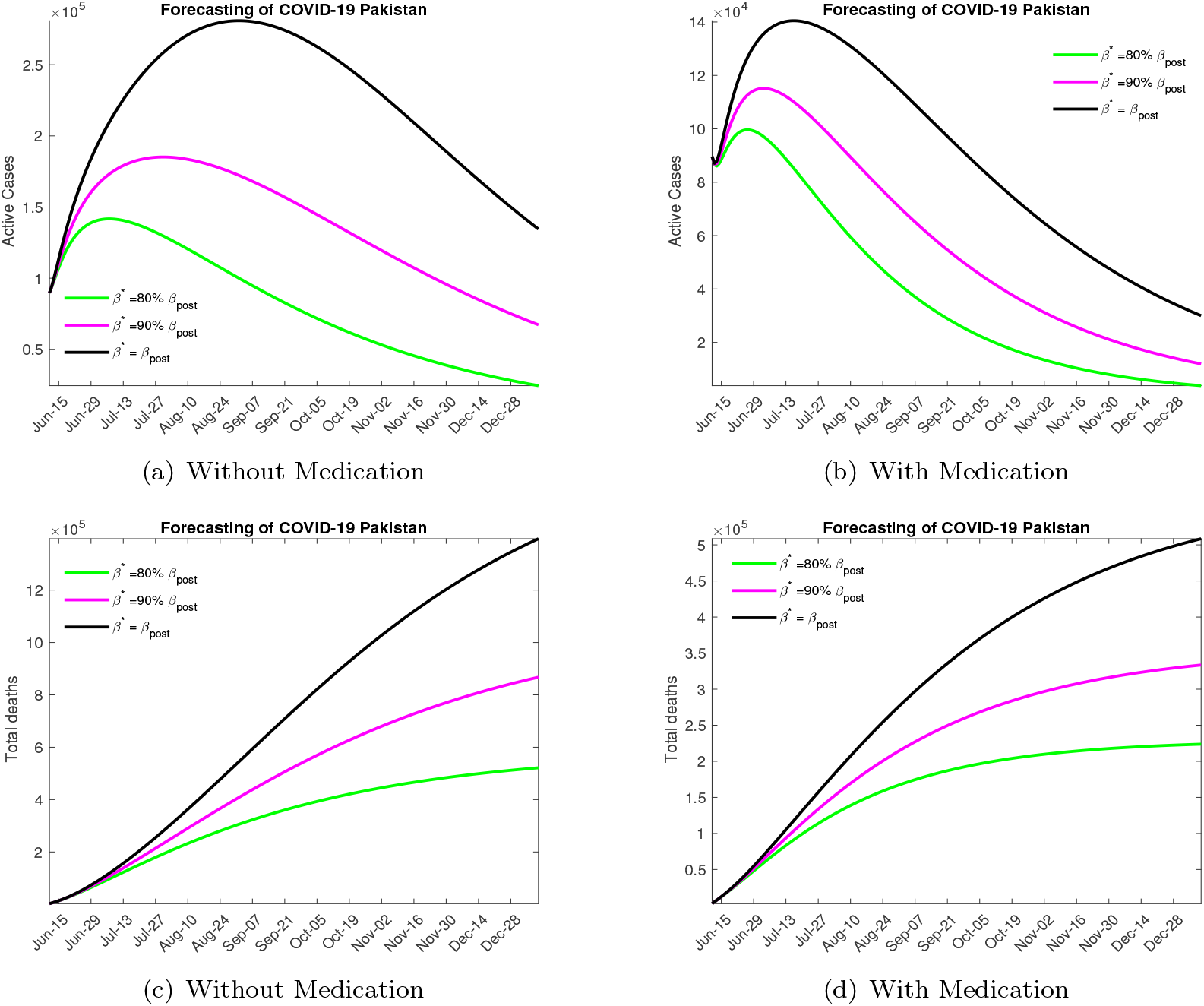
Forecasting with Moderate Lock down

In our opinion the most likely scenario to be played out over the next few month will involve moderate lock down and high social distancing efforts.

In Figure 7 above we consider the situation when a strict lock down is imposed increasing the quarantine rate to pre lock down level. Without medication Figure 7 (a) the peak will be reached in early to mid July (depending on the contact rate) with the maximum number of active infected being around 125000 *−* 200000. In this scenario 300000 *−* 700000 disease related deaths are expected by the end of the year. In Figure 7(b) when medication is available the disease peaks between late June to early July with the maximum active infected number being around 90000 *−* 125000 (again depending on the social distancing measures), with around 150000 *−* 300000 deaths due the the infection.

**Figure 7:**
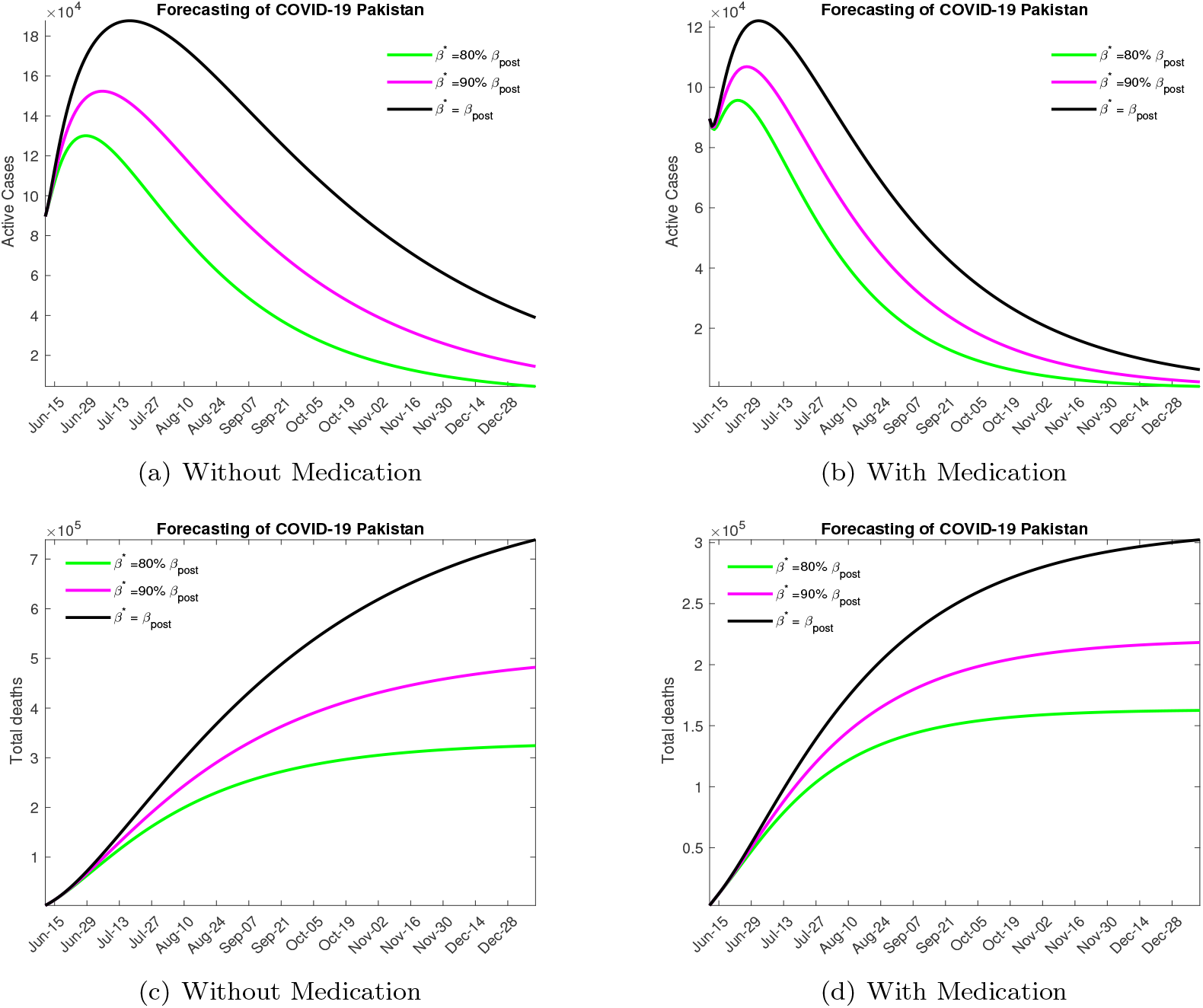
Forecasting with Strict Lock down

We would like to point out that this level of quarantine is not very realistic, considering the economic and social costs the government is unlikely to enforce such stringent lock down measures.

## 6 Conclusion

In this study we have estimated the severity of the COVID 19 outbreak in Pakistan, modelling the transmission dynamics of the disease using an extension of the SEIR model. The model includes important transmission pathways such as asymptomatics, quarantined and isolated individuals as well as the medicated population sub group. We also use this model to project the epidemic curve and give estimates of the disease burden under various realistic control strategies.

An ordinary differential equation based model is used for the transmission dynamic of COVID 19 as described above. Disease free equilibrium (DFE) and an en equilibrium when the disease is endemic in the population is derived. We establish a threshold quantity ℛ_0_ in terms of the parameters of the model, observing that the disease dies out whenever ℛ_0_ *<* 1 and is endemic if ℛ_0_ *>* 1. We study the variation of ℛ_0_ on some of the model parameters, specifically on the parameters which can be varied by various control mechanisms, such as quarantine, social distancing and medication. We note that quarantine and social distancing and medication can help in lowering the value of ℛ_0_ and bring the epidemic under control, with quarantine being the most effective mechanism followed by social distancing and medication. However there are practical difficulties in establishing a strict quarantine as is evident from the relaxation of lock downs in most countries, hence a mix of all these strategies is perhaps the best way forward.

Noting that in Pakistan the epidemic progressed in two distinct phases, before and after the lock down restrictions were relaxed, we estimate the value of ℛ_0_ separately for both these phases. For the pre relaxation phase we estimate ℛ_0_ = 1.07 and for the post relaxation phase our estimate comes out to be ℛ_0_ = 1.86. This is consistent with the growth dynamics and other estimates in the literature.

These results are then used to project the epidemic curve. We explore various possible scenarios, with varying levels of lock down, social distancing and medication. The lock down and social distancing measures are captured by quarantine rate (*ϵ*) and contact rate (*β*) in our study. In the case where a strict lock down is enforced taking the quarantine rate back to pre-relaxation level, varying the contact and medication rates, we observe that in the best case scenario with high level of all controls the epidemic will peak around late June to early July with a maximum active infected number around 90000. For a moderate lock down which is modelled by taking the quarantine rate to be twice the post-relaxation rate, and varying the contact and medication rates we note that the epidemic will peak around mid July with the active infected numbers around 120000 at that point considering that social distancing measures are enhanced. Finally, we look at the case when no further lock down measures are taken and the quarantine rate remains at the value estimated on June 15, in this scenario we note that the epidemic will peak around late September with the maximum active infected number around 700000.

In this work we estimated the severity of the COVID 19 outbreak in Pakistan, noting that the epidemic followed two different trajectories before and after lock down restrictions were relaxed. The restrictions we observe kept the value of ℛ _0_ close to 1, however once the restrictions were eased the number of cases increased at a high rate, reflected in ℛ _0_ = 1.86. We also generated possible future trajectories based on intervention and control measures to varying degrees. In case no significant measures are taken at the peak of the epidemic, which will occur in September the maximum active cases will cross the 700000 mark while in a very optimistic scenario this will happen in early July with maximum active cases contained to around 150000. We strongly agree with the recent WHO recommendation that some form of moderate lock down needs to be reinforced, this along with social distancing and possible medication will result in a disease peak sometime in mid July and contain the maximum active infected numbers to around 120000. Inaction at this point of the epidemic is projected to have very serious consequences both in terms of disease burden and mortality.

## Data Availability

https://github.com/CSSEGISandData/COVID-19

## 7 Appendix

### 7.1 Proof of Lemma 3.1

*Proof*. Adding the equations of model (2),this results in change in total population as

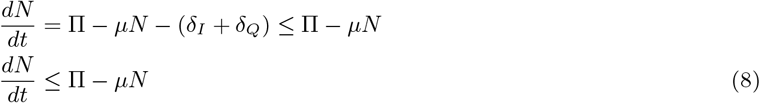

It follows that

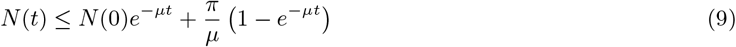

Thus model (2) solutions exists for given initial conditions, and are eventually bounded on every finite time interval.

□

### 7.2 Proof of Lemma 3.2

*Proof*. Using (8) and (9), it follows that as time approaches to infinity *t → ∞*, the population is bounded by the positive number so the set 𝒟

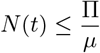

therefore, the set 𝒟 is positively invariant.

□

### 7.3 Proof of Lemma 4.2

*Proof*. Consider the Lyapunov function for model (2).

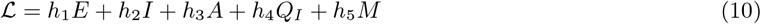

where

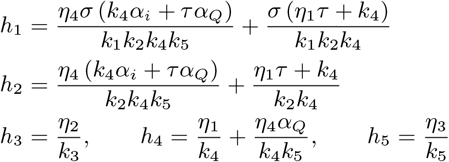

The Lyaponov derivative 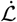 is given as

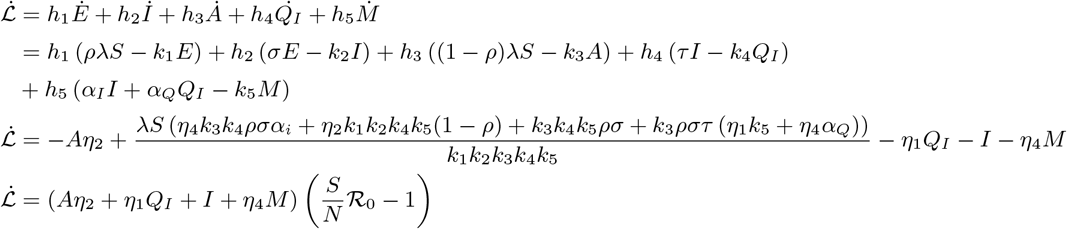

Since *S*(*t*) *≤ N ∈* 𝒟, Thus

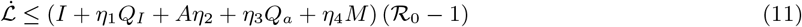

Thus, if 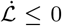 if ℛ_0_ *≤* 1 with 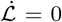 iff *Q*_*s*_ = 0, *E* = 0, *Q*_*E*_ = 0, *I* = 0, *A* = 0, *Q*_*A*_ = 0, *Q*_*I*_ = 0 and *M* = 0. Additionally, the super compact set 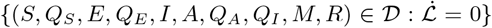 is the singleton ℱ_0_. Thus, LaSalle Invariance Principle Theorem 6.4 [28] guaranties that every solution to the model (2) initial conditions from 𝒟 converge to DFE ℱ_0_ as *t → ∞*. Hence, (*Q*_*s*_, *E, Q*_*E*_, *I, A, Q*_*A*_, *Q*_*I*_, *M*) *→* (0, 0, 0, 0, 0, 0, 0, 0) as *t → ∞*, It follows that *{*(*S, Q*_*S*_, *E, Q*_*E*_, *I, A, Q*_*A*_, *Q*_*I*_, *M, R*)*} →* (*S*^*\**^, 0, 0, 0, 0, 0, 0) as *t → ∞* for ℛ_0_. Hence, ℱ_0_ is *GAS ∈* 𝒟 for ℛ_0_.

**Table 2:** Description of the variables of the model (2)

| Variable | Description |
| --- | --- |
| $N$ | Total population |
| $S$ | Susceptible individuals |
| $Q_S$ | Quarantine of susceptible Individuals |
| $E$ | Individuals Exposed to corona virus |
| $I$ | Individuals Infected with corona virus |
| $A$ | Asymptomatic Individuals |
| $Q_I$ | Isolated individuals |
| $M$ | Medication of individuals |
| $R$ | Recovered from corona virus |

**Table 3:** Description of the parameters of the model

| Parameters | Description | Values |  |
| --- | --- | --- | --- |
| $\Pi$ | Recruitment rate of humans | 100 | Assumed |
| $\mu$ | Natural death rate of humans | 60 years | Assumed |
| $\delta_I$ | Disease-induced death rate individuals | $0.02 \text{ day}^{-1}$ | Estimated from data |
| $\delta_Q$ | Disease-induced death rate of isolated individuals | $0.02 \text{ day}^{-1}$ | Estimated from data |
| $\frac{1}{\sigma}$ | Incubation rate of susceptible individuals | 12 days | Estimated from data |
| $\frac{1}{\gamma}$ | Recovery rate of infected individuals | 4 days | [23, 29] |
| $\beta$ | Effective contact rate | 1.7 | Estimated from data |
| $\rho$ | Probability of getting exposed to covid-19 | $70\% = 0.7$ | Estimated from data |
| $\tau$ | Isolation rate of infected individuals | 0.1 | [23] |
| $\alpha_I$ | medication rate of infected individuals | 0.1 | Assumed |
| $\theta_A$ | Recovery rate of Asymptomatic individuals | 0.1 | Estimated from data |
| $\theta$ | Recovery rate of Isolated individuals | 0.1875 | [23, 29] |
| $\theta_M$ | Recovery rate of medicated individuals | 0.375 | Assumed |
| $\epsilon$ | susceptible Quarantine rate of Susceptible individuals | 0.08 | Estimated from data |
| $\xi$ | Waning rate of susceptible quarantined individuals to susceptible class | 28 Days | Assumed |
| $\eta_1$ | Modification parameter for relative infectiousness of Isolated individuals | 0.4 | Assumed |
| $\eta_2$ | Modification parameter for relative infectiousness of asymptomatic individuals | 0.5 | [23] |
| $\eta_4$ | Modification parameter for relative infectiousness of medication | 0.5 | Assumed |

